# Noninvasive respiratory support in acute hypoxemic respiratory failure associated with COVID-19 and other viral infections

**DOI:** 10.1101/2020.05.24.20111013

**Authors:** Claudia Crimi, Alberto Noto, Andrea Cortegiani, Pietro Impellizzeri, Mark Elliott, Nicolino Ambrosino, Cesare Gregoretti

## Abstract

**Introduction:** Noninvasive respiratory support (NRS) such as noninvasive ventilation (NIV) and high flow nasal therapy (HFNT) have been used in the treatment of acute hypoxemic respiratory failure (AHRF) related to the coronavirus disease (COVID-19) and other viral infections. However, there is a lack of consensus in favor of or against NRS use due to the risks of worsening hypoxemia, intubation delay, and aerosols environmental contamination associated with the use of these tools. We aimed to summarize the evidence on the use of NRS in adult patients with COVID-19 and other viral pneumonia (i.e. H1N1, SARS, MERS) and AHRF. We also searched for studies evaluating the risk of aerosolization/contamination with these tools.

**Evidence Acquisition:** We searched MEDLINE, PubMed EMBASE and two major preprint servers (biorXiv and medRxiv) from inception to April 14, 2020, for studies on the use of respiratory support in AHRF and viral pneumonia.

**Evidence Synthesis:** The search identified 4086 records and we found only one randomized controlled trial out of 58 studies included, with great variabilities in support utilization and failure rates. Fifteen studies explored the issue of aerosolization/contamination showing a high risk of airborne transmission via droplets generation during the use of these modalities

**Conclusions:** Use of NRS and treatment failure in the context of COVID-19 and viral infection associated-AHRF, varied widely. Dispersion of exhaled air is different depending on the type of respiratory therapies and interfaces. Data from randomized controlled trials are lacking.

## INTRODUCTION

Viruses are the most common causes of respiratory infections and are responsible for pneumonia in over 20% of cases ^1^. Lately, new viruses associated with recent outbreaks have been discovered, such as a new strain of coronavirus, SARS-CoV-2 ^2^.

In the early months of 2020, coronavirus disease 2019 (COVID-19) rapidly became a public health emergency requiring hospitalization in approximately 14% of infected patients ^2^, and it is associated with severe acute hypoxemic respiratory failure (AHRF) in nearly 30% of hospitalized patients requiring oxygen and noninvasive respiratory support (NRS) and intensive care unit (ICU) admission in approximately 5% of cases ^2-4^.

COVID-19 has spread rapidly, overwhelming hospitals and facing the challenging situation that the demand for ICU beds outstrip capacity worldwide. Therefore, NRS such as noninvasive ventilation (NIV), continuous positive airway pressure (CPAP) and high flow nasal therapy (HFNT) represent an essential component of the global response to this emerging infection since it may be performed outside the ICU ^5^, limiting the need for critical care resources and invasive ventilation (IMV). Wang and coworkers ^4^ showed that over 50% of patients with AHRF due to SARS-CoV-2 infection received NRS in China. Yet, the Food and Drug Administration has recently provided guidance that allows hospitals to modify home respiratory devices, to ensure the availability of the greatest possible number of systems for delivering NRS to the highest number of patients during the public health emergency ^6^.

Nevertheless, the beneficial effects of applying NRS in patients with viral pneumonia associated-AHRF remain controversial ^7, 8^, due to the possible rapid deterioration of hypoxemia and the higher rate of endotracheal intubation (ETI) ^9^; furthermore, its safety in diseases transmittable via respiratory route is unclear.

We conducted a review to summarize the clinical evidence on the use of NRS in viral pneumonia and to explore the risk of environmental contamination in healthcare settings.

## METHODS

### Literature search

We conducted searches in Medline, PubMed, and EMBASE for published randomized control trials (RCTs) and observational studies (both prospective and retrospective) in adult patients from inception to April 14, 2020, in English only.

Searches were performed using a combination of the following terms: *COVID-19, virus infections, noninvasive respiratory support, NIV, BiPAP, CPAP, HFNT*, and *pneumonia*. We also considered articles evaluating aerosolization into the environment during respiratory therapies.

To increase the sensitivity, we searched in preprint servers (biorXiv and medRxiv) for COVID-19 and SARS-Cov-2 articles ^10-13^. Additional relevant articles were identified through bibliography of the retrieved papers (“snowballing method”). Detailed search strategy was reported in the **Supplementary material 1**.

Two investigators (CC, AN) independently screened the titles and the abstracts retrieved, and selected full-text of relevant articles, (**Supplementary material 2**). Disagreements between reviewers were discussed and a third author (AC) was consulted if agreement could not be reached.

## RESULTS

The literature search identified 3089 records and 997 articles from preprint servers (780 medRxiv, 217 bioRxiv). Review of titles and abstracts retrieved 404 potentially relevant full-texts with 4 from the preprint server. After full-text analysis 8 articles were added from snowballing and 354 articles were excluded for the following reasons: 191 focused on pediatric populations, 22 focused on pneumonia in immunocompromized patients, 49 were case reports, 40 were abstract/conference papers, 14 editorial/letter, 13 reviews, 10 not available, 15 not relevant. A total of 58 articles were reviewed, (**Supplementary material 2**). The review was conducted in accordance to PRISMA statement..

### COVID-19

COVID-19 is a recent and still ongoing pandemic that has spread quickly, overwhelming hospitals, increasing the demand for critical care services with a mortality rate in patients requiring advanced respiratory support ranging from 62 to 97% ^14-16^, with nearly 25% of patients remaining critically ill ^17^. In this context, an increasing number of publications are becoming available on the use of NRS ^3, 4 10-13, 16-21^, mostly from China ^3, 4 10-13, 16, 18, 20, 21^, but also from Italy ^19^ and USA ^17^.

The percentage of patients requiring NRS varied greatly among the studies, ranging from 11% to 96%, (Figure 1), with higher usage rate in China (62% on average), and lower rate in North America (20%) ^17^ and Italy (11%) ^19^, with Italian authors reporting a higher need for IMV in their critically ill population compared with other studies ^20, 21^.

**Figure 1.**
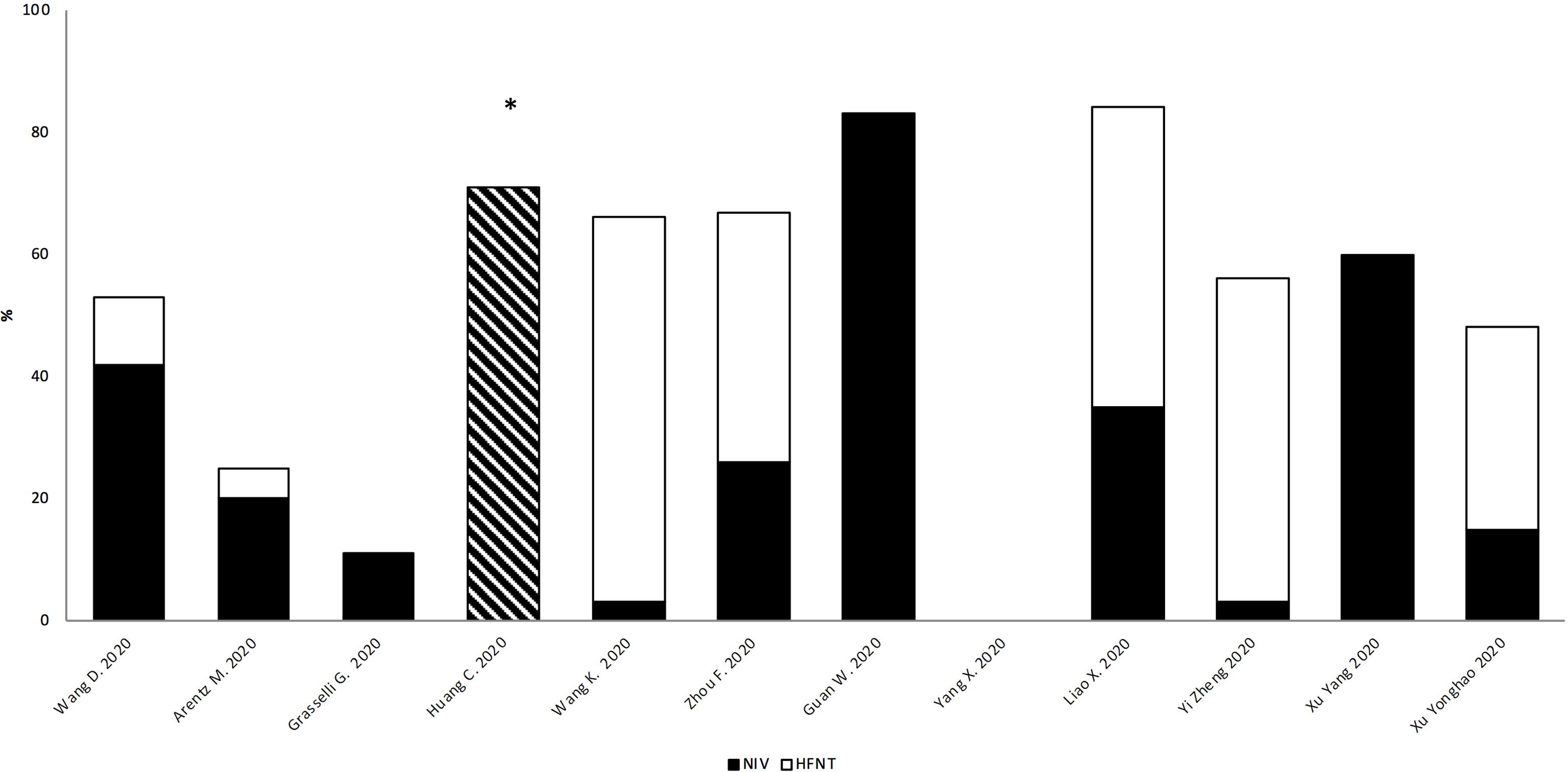
Noninvasive respiratory supports utilization rate among COVID-19 studies. Each bar indicates the total percentage of noninvasive supports usage, black and white bars indicate proportions between NIV and HFNT. *Huang C: only total data available with no distinction between NIV and HFNT NIV: noninvasive ventilation; HFNT: High flow nasal therapy.

Noninvasive support was mainly delivered in ICU, apart from two studies that described its use outside the ICU settings in 20% ^3^ and 50% of their population ^20^, respectively. The overall mean utilization rate among studies was 31% for HFNT ^4, 10, 12, 13, 17, 18, 20^ and 30% for NIV ^4, 10-13, 17-21^, with Wang et al. ^18^ reporting a prevalent use of HFNT (63%) as initial respiratory support since that, because of the emergency, the clinical staff involved in patients’ management had a heterogeneous background and little or no experience with NIV.

Failure rate was mainly explored in Chinese studies and varies widely as shown in the **Supplementary material S3**, ranging from 52% ^21^ to 92% ^20^: however, data are not uniform since sometimes NIV was used as a rescue therapy after HFNT failure ^18^.

Mortality rate was rarely reported in the studies included. Yang at al. ^16^ in their cohort of 52 critically ill patients showed a 48% mortality rate (16/33) for patients receiving HFNT and 72% (23/26) for patients treated with NIV as the first-line treatment or as an escalation of care.

### Influenza A-H1N1

Influenza A (H1N1), also called swine flu, spread globally at the beginning of 2009, starting from Mexico and causing a febrile respiratory infection, reporting mortality of more than 50% in mechanically ventilated patients ^22^.

As shown in Figure 2, a high variability (from 13 to 80%) in NRS utilization rate was found among patients requiring ventilatory assistance, with wide variations in success rate reported within similar geographical areas ^23-27^.

**Figure 2.**
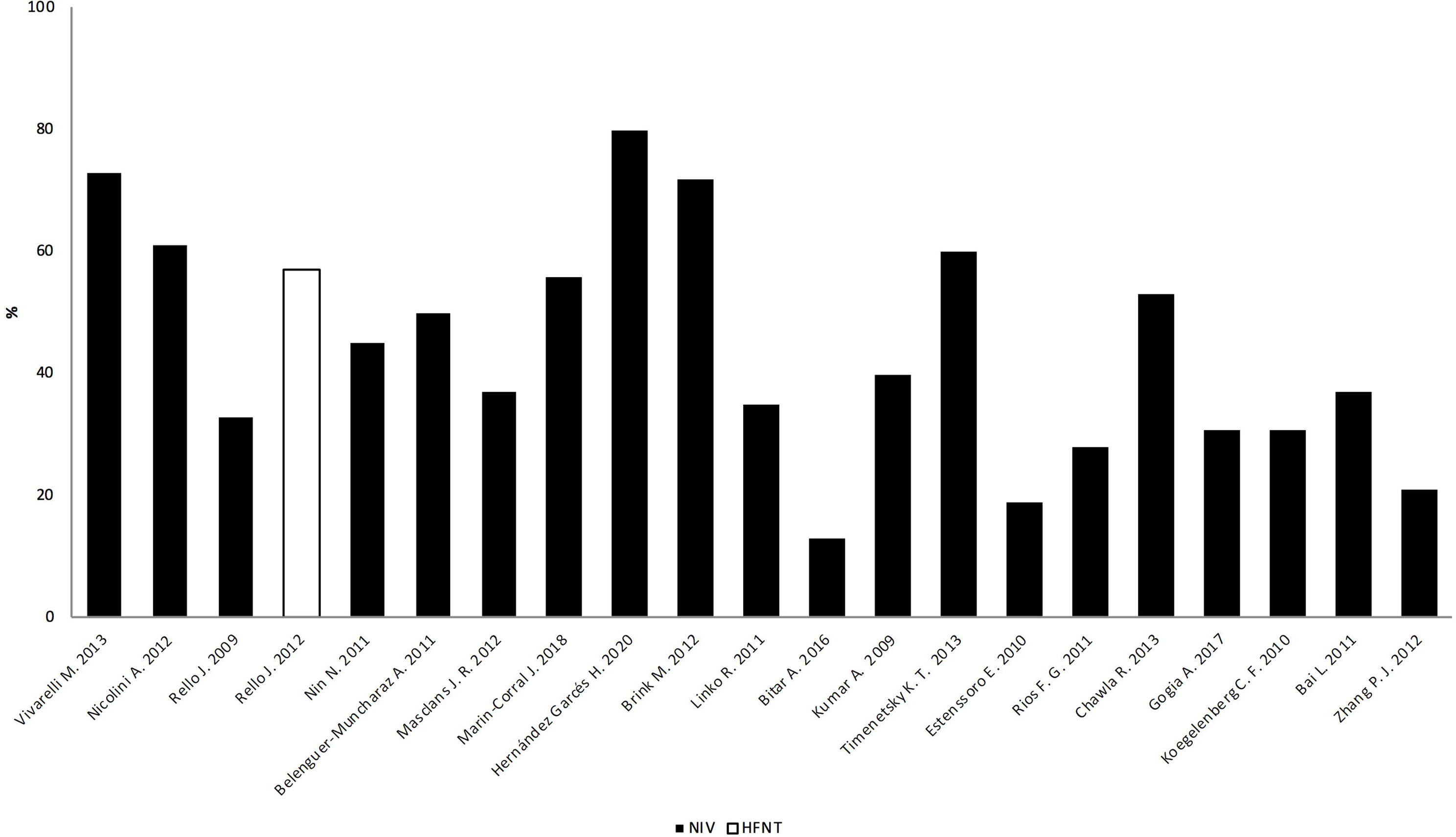
Noninvasive respiratory supports utilization rate among H1N1 influenza studies. Each bar indicates the total percentage of noninvasive supports usage, black and white bar indicate proportions between NIV and HFNT. NIV: noninvasive ventilation; HFNT: High flow nasal therapy.

Failure rate was very heterogeneous among the studies as shown in Table 1 and **Supplementary material S4**. This wide variability is caused by the different analytical methods used in each study: type of enrolled patient populations, sample size and criteria for ETI.

**Table 1.**
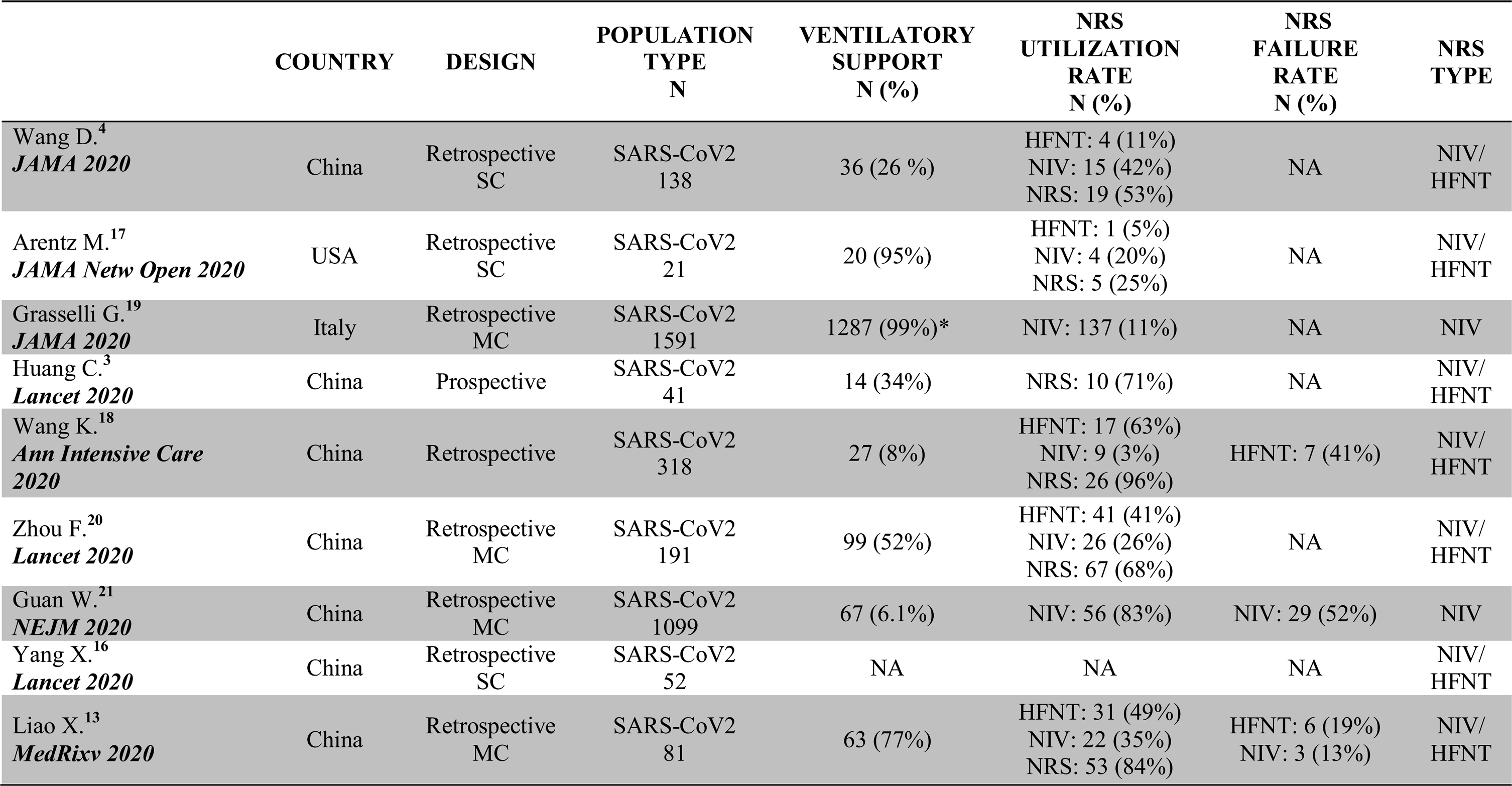
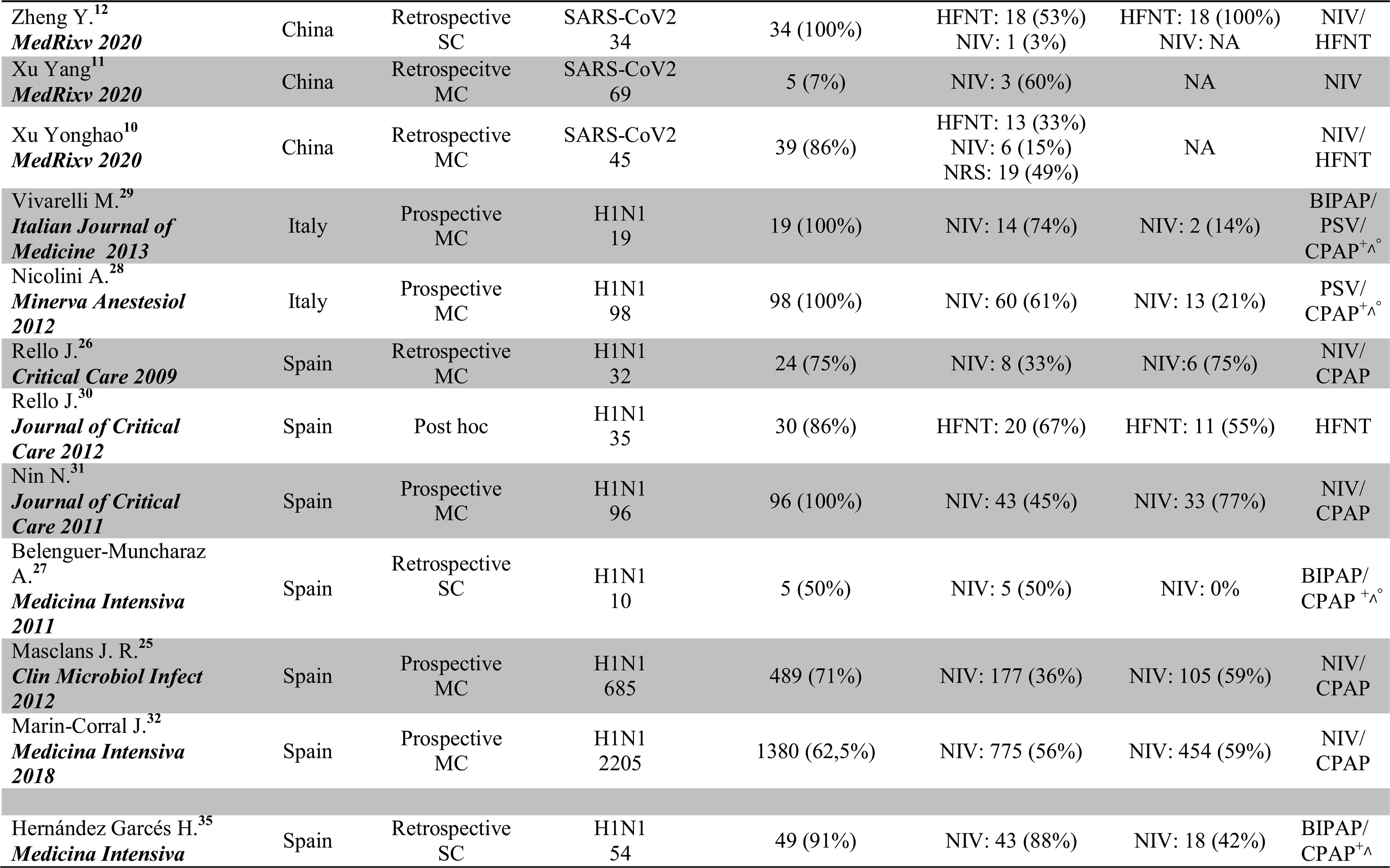
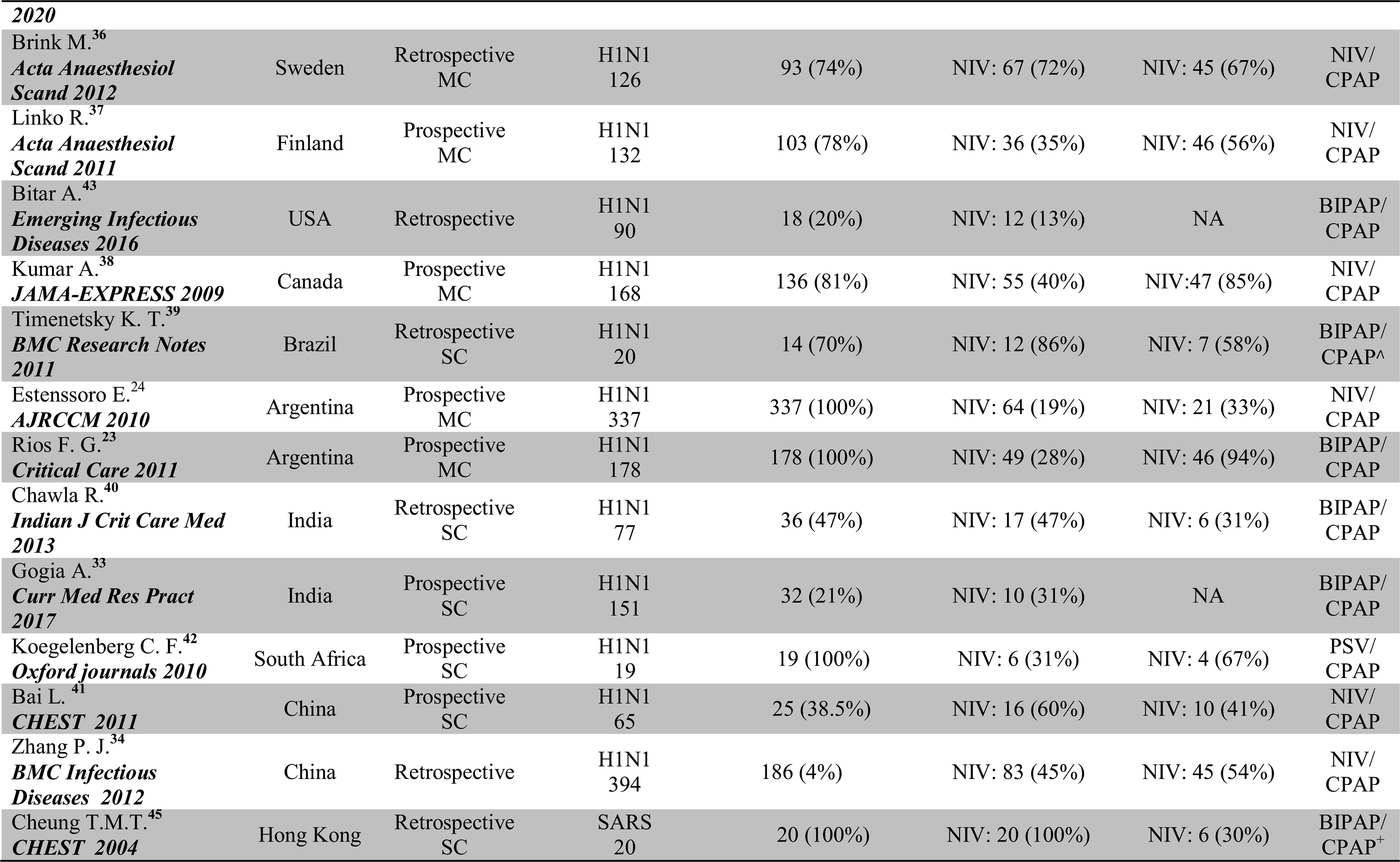
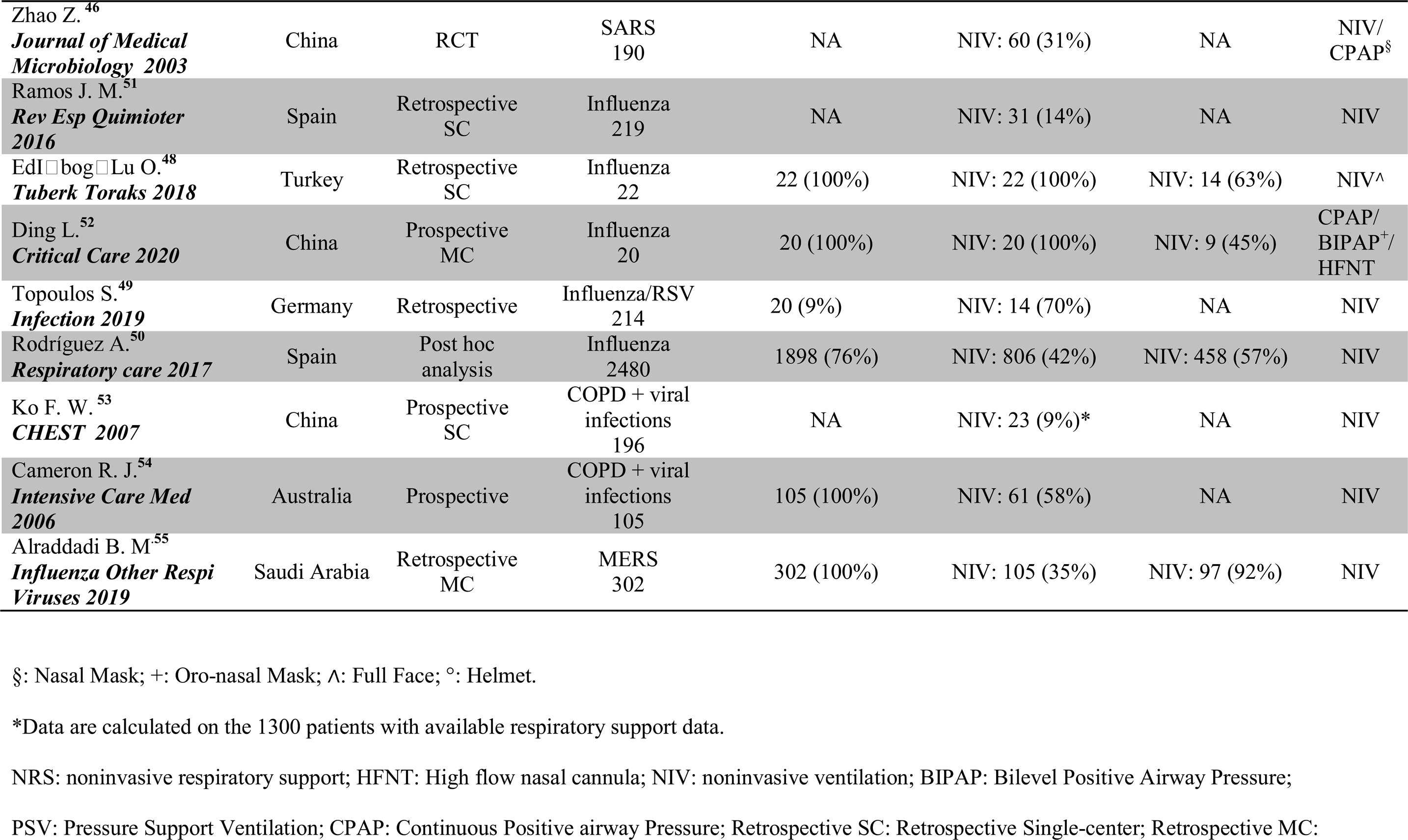
Characteristics and outcomes of included studies.

Few studies ^25, 28-30^ reported the specific criteria before NRS initiation. Almost all the studies were conducted in ICU settings ^23-27, 30-42^ except two that were performed in both ICUs and High Dependency Units (HDUs) ^28, 29^.

Particularly, modes and settings of NRS delivery were rarely described ^23, 27-30, 35, 39, 43^. Interfaces used were specified in only 6 out of 21 studies (28%) ^27-30, 35, 39^. Oro-nasal and total face masks ^28, 29, 35, 39^ were the most widely used interfaces for NIV/CPAP, with the use of the helmet reported only in the Spanish experience ^27^, as shown in Table 1.

Three studies highlighted predictors of success ^25, 29, 37^. Factors associated with a positive response to NIV treatment were explored by Masclans et al. ^25^ [less radiological involvement, hemodynamic stability and a sequential organ failure assessment (SOFA) score < 7]; similar results [fewer pulmonary lobes involved, lower Simplified Acute Physiology Score (SAPS II) and SOFA score, higher arterial oxygen tension to inspiratory oxygen fraction (P/F)] were showed by Vivarelli et al. ^29^ and by Linko et al. ^37^.

Predictors of failure were highlighted in three studies ^26, 28, 41^. Rello and coworkers ^30^ showed that HFNT failure was associated with a SOFA score value of 4 or more, Acute Physiology And Chronic Health Evaluation II score (APACHE II) of 12 or more and shock; Bai et al. ^41^ identified a higher APACHE II scores on presentation as associated with unfavorable NIV outcome, while Nicolini and coworkers ^28^ found that high SAPS II score, P/F ratio <127 and the new infectious complications related to the ICU stay were determinants for NIV failure.

Data on mortality are very heterogeneous among the studies and only six authors described a specific NIV failure/success-related mortality ^25, 28, 36, 39, 40, 42^ with no changes in terms of mortality compared to patients who were directly intubated in two studies ^36, 40^.

### Severe Acute Respiratory Syndrome (SARS)

SARS-CoV was identified in 2003 in Hong Kong and caused AHRF in nearly 25% of affected patients ^44^.

In a retrospective analysis ^45^, NIV was effective in preventing ETI in 70% of patients (14/20) with a low APACHE score. In a study ^46^ with a random assignation to 4 treatments, the best response (no deaths) was seen in the group of 60 patients receiving early high-dose steroids and nasal CPAP. Failure rates are shown in **Supplementary material 5**.

### Influenza

Seasonal influenza epidemics may be associated with high morbidity and mortality, especially in older patients, since symptoms may progress quickly to respiratory distress and extensive pulmonary involvement ^47^.

In an analysis of 22 patients admitted to ICU for influenza-related AHRF in Turkey, all patients were treated with NIV with a full face mask as a first-line intervention ^48^ with a survival rate of 78% in patients where NIV was successful.

In a report ^49^ of 651 patients with influenza and respiratory syncytial virus, 32% (61/191) developed pneumonia and 33% of them required ventilatory support, predominantly noninvasive (70%). A secondary analysis from a large prospective observational multi-center study ^50^ of 1898 critically ill subjects admitted to the ICU comparing patients who received NIV or IMV immediately after ICU admission showed that NIV was the treatment of choice in 42.4% of them, with 56.8% treatment failure rate in more severe patients with higher APACHE II and SOFA scores and more pronounced radiological infiltrates, and higher mortality (38%). Patients with a SOFA score ≥ 5 had a 3-fold risk of NIV failure; moreover, a higher NIV success rate was reported in patients with chronic obstructive pulmonary disease (COPD), (73.7%).

A retrospective Spanish study ^51^ of 219 hospitalized patients with AHRF showed that patients older than 80 years old were more likely to receive NIV treatment as compared to younger patients (22% vs 9.3%).

A recent observational study by Ding et al. ^52^ on patients with moderate-to-severe ARDS (SpO2>95%) showed the possibility of successfully managing patients with early HFNT combined with prone positioning or NIV alone, improving oxygenation and avoiding intubation in nearly 55% of the patients (11/20).

Finally, analyzing viral-related COPD exacerbation, a study ^53^ of 76 patients with acute exacerbations of COPD showed that NIV was successfully used in nearly 23 episodes (8.8%). In another study on 105 COPD patients with viral exacerbation, 61 patients (57%) received NIV alone, with 17 (16%) receiving both NIV and ETI and only 29 (27%) being intubated without NIV ^54^.

### Middle East respiratory syndrome (MERS)

The use of NIV has also been studied in 302 critically ill patients with MERS ^55^ where NIV was provided in 105 (35%) patients, presenting with a lower baseline SOFA score and less extensive infiltrates on chest radiograph compared with patients managed with IMV with similar ICU and hospital length of stay. The intubation rate in the NIV group was 92.4%, with a 90-day mortality of 65.7% vs. 76.1% of the IMV group; the use of NIV was not independently associated with mortality. Failure rates are shown in **Supplementary material 5**.

### Viral aerosol droplet generation

Biohazardous aerosols are routinely generated in hospitals by processes such as breathing, coughing, sneezing, ETI, NIV, delivery of medication by nebulization. Aerosol generating procedures may expose health care workers to bacterial/viral pathogens causing acute respiratory infections. Some procedures (ETI, NIV, tracheotomy, and bag ventilation) are believed to spread more aerosols and droplets ^56^.

Currently, droplets are defined as being >5 μm in diameter that fall rapidly to the ground under gravity and therefore are transmitted only over a limited distance (e.g. ≤1 m). Airborne transmission is different from droplet transmission as it refers to the presence of microbes within droplet nuclei, which are generally considered as being particles <5μm in diameter that can remain in the air for long periods and be transmitted to others over distances greater than 1 m ^57^.

Noninvasive ventilation is a large (10 μm) droplet (not aerosol)-generating procedure ^58^. Due to their large mass, most fall out on to local surfaces within 1 m. Droplets’ productions during NIV are greatly reduced with the use of non-vented full-face masks connected with a viral/bacterial filter ^58^, Figure 4. We identified 15 studies evaluating aerosolization during NRS.

**Figure 3.**
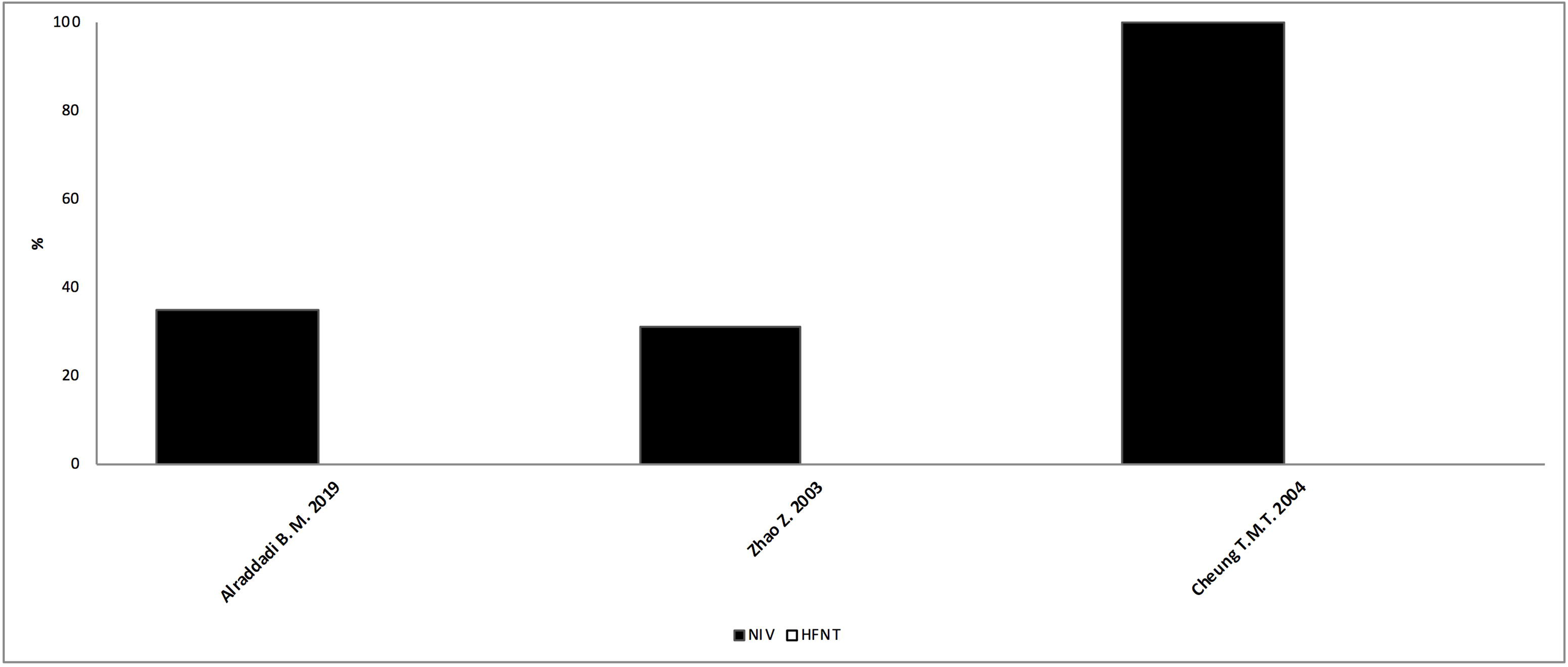
Noninvasive respiratory supports utilization rate among SARS/MERS studies. Each bar indicates the total percentage of noninvasive supports usage, black and white bar indicate proportions between NIV and HFNT. NIV: noninvasive ventilation; HFNT: High flow nasal therapy; SARS: Severe Acute Respiratory Syndrome; MERS: Middle East respiratory syndrome.

**Figure 4.**
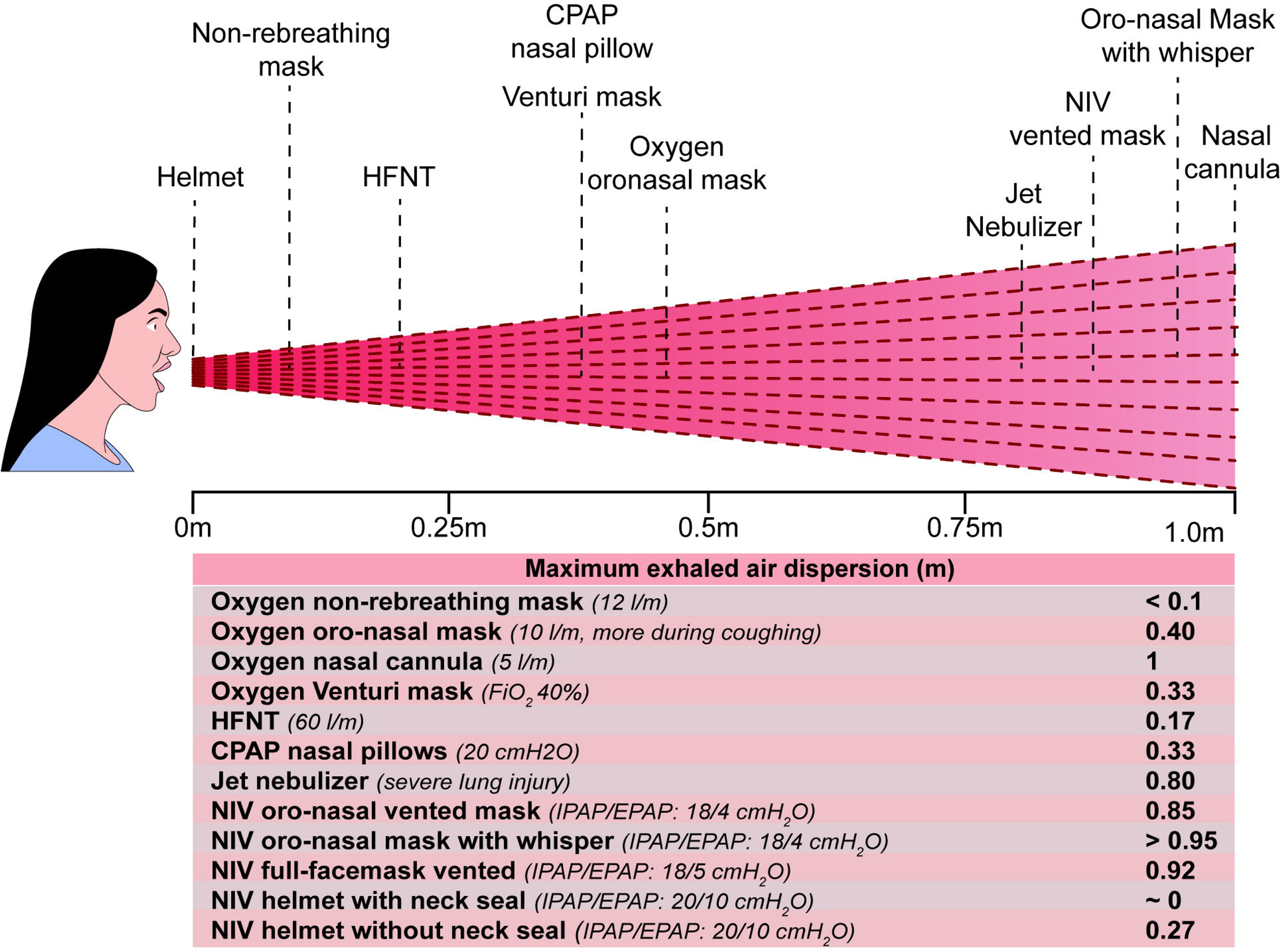
Droplets aerosolization distance. NIV: noninvasive ventilation; HFNT: High flow nasal therapy; CPAP: continuous positive airway pressure.

Hui et al. ^59^ measured airflow using a smoke-marker and confirmed the difference between vented and non-vented masks, measuring the maximum exhaled air distances with different oxygen therapy devices: nasal cannula, Venturi mask, and a non-rebreathing mask, Figure 4. Authors also recommended that a Whisper swivel valve would not be advisable in managing patients with febrile respiratory illness of unknown etiology ^60^, especially in the setting of an influenza pandemic, even if the addition of a viral filter before the expiratory port can reduce aerosol emission. It is also important to avoid the use of higher inspiratory positive airway pressures, which could lead to a wider distribution of exhaled air and possible room contamination ^60, 61^. It has been demonstrated that the helmet is the preferred NIV interface to reduce leakage and droplet contamination (NIV configuration with double limb circuit) especially the model provided with a neck seal. A helmet without a good seal at the neck can produce a radial leakage of 270mm ^62^.

The leakage produced during procedures and ventilation strategy led to different room contamination accordingly to the room dimensions (i.e. more contamination in smaller rooms) ^63^. The clinical usefulness of NRS in AHRF is still debated, but moreover, in the context of viral pneumonia, it is important to understand how a flow up to 60 l/m impact on the spread of droplets. Recent data shows that HFNT did not increase the spread of exhaled air despite operating at higher flow rates, possibly because of the very low positive end-expiratory pressure (PEEP) generated ^64^. Another study, not based on simulation, has shown that HFNT did not increase bacterial contamination (airborne and surface) compared to an oxygen mask ^65^.

Most researches were based on the smoke visualization of the exhaled air and not on the direct measurements of the viral droplets.with the exceptions of only two studies ^58, 65^. Therefore, we can consider it as the maximum boundaries and trajectory of the droplets dispersion. Further studies with direct measurements of pathogen agents are required to plan healthcare protection during viral infection.

## DISCUSSION

To the best of our knowledge, this is the most up to date systematic review evaluating the use of NRS in the treatment of AHRF related to viral infections. We identified a large body of literature that describes current clinical practices and challenges of noninvasively treating those patients. However, most of the studies had a non-randomized design, so uncertainty remains on the safety and effectiveness of NRS use in this setting. Our results should encourage the scientific community to promote future research in this area to fill the knowledge gap, especially in the current context of the COVID-19 pandemic.

There is a theoretical strong rationale for the use of NRS as compare to standard oxygen in patients with pulmonary infiltrates. The application of PEEP through NIV/CPAP can effectively increase functional residual capacity and re-inflate collapsed alveoli, improving ventilation/perfusion matching ^66^. High-flow nasal cannula generates a small PEEP effect, improving oxygenation, respiratory mechanics and end-expiratory lung volume, reducing the work of breathing and offering good comfort to the patient ^8, 67^.

Furthermore, physicians are likely to use both techniques in clinical practice. In a recent survey of 235 ICU physicians in France ^68^, HFNT and NIV use was perceived as relevant in managing AHRF by all the participants and in a previous European survey on NIV practices ^69^ physicians reported the use of NIV for *de novo* ARF in about 15-20% of patients, with intensivists more likely to use it than pulmonologists.

Nevertheless, clinical benefits are less clear and guidelines ^7^ did not provide any recommendation in favor or against NIV use in both *de novo* and viral pandemic AHRF because the published literature so far did not identify a clear advantage in terms of outcomes ^70^; similar conflicting evidence exists on the use of HFNT as well ^8, 71^. However, during the H1N1 pandemic in 2009, a statement of international societies ^72^ suggested that NIV should not be used routinely as an alternative to IMV but it might be applied in highly selected cases to prevent progression of AHRF and need for ETI or as “ceiling” treatment during pandemics.

In the examined studies we found wide variability in terms of utilization rate, timing of initiation, operational mode and settings, types of interface, where to perform it and, cut-off parameters for withdrawal. The results of this review provide an outline of common themes for future research and judicious reflections: what might be the role of NRS in viral pneumonia AHRF? We believe that the following should be considered.

First, the utilization rate in viral pneumonia-associated AHRF is not uniform among the evaluated studies, ranging from 11% to 96% during the COVID-19 pandemic. Among noninvasive devices, HFNT seems more used than NIV ^10, 12, 13, 18, 20^ in the COVID-19 scenario except in 2 studies ^4, 17^, probably because it requires less expertise ^18^ and it can be easily used also outside the ICU settings ^5^. Little information on success or failure rate is available ^18, 21^ with a reported overall mortality of patients requiring advanced respiratory support of about 63% ^14^ or higher ^16^. Similar fluctuating usage rates were reported during H1N1 and MERS/SARS outbreaks ranging from 13% to 80% and from 35 to 100%, respectively, with NIV predominantly used since HFNT was released in 2009. Moreover, we found striking differences in the reported success rate, going from encouraging ^24, 28, 45^ to concerning ^36, 38, 55^, highlighting the need for performing NRS only in highly expert centers ^28^ and with well-defined patient selection and ETI criteria.

Second, timing and patient selection should be clearly defined. Most of the examined studies showing benefits of NIV suggested the early enrollment (P/F<300) ^28^, or to start in subjects with less severe disease ^25, 29, 30, 55^, even if these patients might have got better anyway. Few studies described criteria of initiation of NRS ^29, 48, 50^; most of the trials are retrospective and sometimes the decision to start was made by the physician on duty on clinical rounds. Based on these findings, the interval between the onset of AHRF and the initiation of a gentle NRS trial is crucial to “buy time” for medical therapy to take effect and possibly reverse the underlying cause of respiratory failure. Likewise, a rapid and wise assessment of slight variations in gas exchange and patient status has to be made during trials to avoid the risk of intubation delay. This is extremely important since if NIV is successful, patients’ mortality is negligible, but if NIV fails, mortality is higher as compared to patients receiving ETI as first-line treatment ^73^. Similarly, patients intubated after HFNT showed greater mortality if experience late failure (>48h, 66% vs. <48h, 39%) ^8^.

Third, modes and settings are heterogeneous and not specified. Almost all the examined studied reported usage but do not describe in-depth modes (i.e. BiLevel vs. CPAP)^3^, settings ^23, 27-30, 35, 39, 43^ (i.e. levels of PEEP or pressure support). This is an important aspect to consider since CPAP may be useful in reversing atelectasis but the inadequate pressure support in patients with high respiratory drive may generate patient self-induced lung injury (P-SILI) ^74^. Furthermore, NIV cannot control tidal volumes and it cannot guarantee lung-protective ventilation in nearly 75% of patients, which might worsen ventilator-induced lung injury ^75^. Very few studies specify the type of interface used ^27-30, 35, 39, 52^. The selection of non-vented interfaces or helmet for delivering NIV/CPAP and the application of a surgical mask when using HFNT is crucial to reduce the risk of viral droplets dispersion with the risk of transmitting the infection to healthcare workers ^65^ that should always use personal protective equipment to protect themselves ^76^. In particular, helmet may confer advantages over the face mask: it can minimize virus spread within treatment facility ^62^, and it can be used without a ventilator connected to fresh gas flow ^77^, even if it has to be considered that some hospitals may have difficulties in coping with the high oxygen flow demand.

Fourth, hospital settings are heterogeneous. Only in two studies, NRS has been performed also in non-ICU settings ^3, 20^. Where NRS is performed in each hospital around the world is not uniform and might depend upon hospitals’ internal organization and total numbers of ICU beds available. An underestimation of the progression and severity of *de novo* AHRF in patients hospitalized in general wards has been shown by Quentin et al. ^78^ who reported that nearly 10% of patients already met the criteria for ARDS but were likely to be transferred to an ICU with a median of 6 days after admission. On the other hand, the ICU might expose the patient to additional risks such as infections that might contribute to unfavorable outcomes ^28^ and it can be excessively costly for patients with mild ARDS, especially during pandemics, where ICU resources should be limited to the sicker patients. Moreover, NIV delivery is an added challenge for the clinician: it is highly time-consuming and needs specific skills to be performed. Probably, to get the most from this noninvasive strategies, specialized care provided by a respiratory and highly expert critical care team ^28^ and an appropriate location such as High Dependency Units (HDUs) ^28, 29^ might be the best compromise to accommodate the major number of patients, deliver cost-effective NRS trials and relieve the burden of ICU beds availability, as it has been described in Italy during the COVID outbreak ^19^.

Finally, predictors of failure or success are lacking. Tools and scores to predict HFNT and NIV failure have been developed. A Rox index ^79^ (SpO2/FiO2)/respiratory rate < 3.85 at 12 hours identified patients at high risk of failure for HFNT trial as well as the HACOR (heart rate, acidosis, consciousness, oxygenation, and respiratory rate) scale > 5 at 1 h for NIV ^80^. However, these tools were not used in the examined studies. Few studies ^25, 26, 28, 35^ identified predictors of failure. None of the studies describe what could be considered an acceptable clinical response or a good time-frame for withdrawal or which is the best strategy to wean the patient (a stepwise reduction of the amount or of the time of support).

We should recognize that most of the NRS studies were inadequately designed to evaluate its role in viral pneumonia. Nevertheless, although RCTs remain the most robust method to study the effects of a medical tool, in specific situations such as pandemics, it might be almost impossible for clinicians to spend efforts in performing properly designed clinical trials since they should focus all available resources on saving lives. Therefore, the studies carried out in real-life especially in this contingency are crucial to confirm the effectiveness of previously studied medical devices.

Probably in the catastrophic scenario of a pandemic, it might appear reasonable to try an early NRS to potentially prevent the progression of AHRF severity and this is likely the reason why in the COVID-19 era, any type of NRS ^81^, even repurposed techniques ^6^ are used to accommodate the major number of patients possibly limiting ICU resources.

## CONCLUSIONS

Despite the lack of robust scientific evidence, NRS is widely used in the treatment of viral pneumonia-related AHRF, with the advantage of reducing the burden of critical care resources utilization but with the risk of delaying but not avoiding intubation.

We found great variability in NRS utilization and treatment failure among different geographic areas and sometimes within the same country. Nevertheless, the published data provided very limited and unclear information about modes, settings, interfaces, locations, timing of initiation, or withdrawal of NRS.

Further high-quality studies are needed in these settings, to explore the efficacy of a system with potential viral droplet spread and possible contamination of health-care workers.

## Data Availability

The authors confirm that the data supporting the findings of this study are available within the article [and/or] its supplementary materials.

## KEY MESSAGES

- Noninvasive respiratory support is widely used in the treatment of viral pneumonia-related acute hypoxemic respiratory failure, despite the lack of solid scientific evidence.
- There is a great variability in noninvasive respiratory support utilization and failure rate and unclear information about modes, settings, interfaces, locations, timing of initiation, or withdrawal.
- There is a lack of randomized control trial that needed to be performed to explore the efficacy of a potential viral droplet generation system such as noninvasive respiratory support in the context of viral infections.

## NOTES

### Authors’ contributions

— CC, AN, conceived the content, drafted the manuscript, approved the final version to be submitted. AC conceived the content, assisted with the literature search strategy, helped in writing the manuscript, approved the final version to be submitted. PI conceived the content, helped in writing the manuscript, approved the final version to be submitted. NA, ME, CG helped in writing the manuscript and revised it critically for important intellectual content, approved the final version to be submitted.

## Acknowledgements

None.

